# Transcriptomic profiling of lymphocytic colitis highlights distinct diarrhoeal pathomechanisms

**DOI:** 10.1101/2023.03.14.23287226

**Authors:** Archana Bhardwaj, Andreas Münch, Julia Montague, Stefan Koch, Philip Rosenstiel, Celia Escudero-Hernández

**Affiliations:** Institute of Clinical Molecular Biology (IKMB), Christian-Albrechts-University and University Hospital Schleswig-Holstein, Campus Kiel, 24105, Kiel, Germany; Department of Biomedical and Clinical Sciences (BKV), Linköping University, Linköping, Sweden; Department of Gastroenterology and Hepatology, Linköping University, Sweden, and Department of Health, Medicine, and Caring Sciences, Linköping University, Linköping, Sweden; Wallenberg Centre for Molecular Medicine (WCMM), Linköping University, Linköping, Sweden

**Author notes:** **CORRESPONDENCE** Celia Escudero Hernández, PhD. This work has been performed at the Dep. of Biomedical and Clinical Sciences (BKV), Linköping University, Ingång 68, plan 13, US campus, 58183, Linköping, Sweden; and at the Lab of Systems Immunology, Institute for Clinical Molecular Biology (IKMB), Christian-Albrechts-University and University Hospital Schleswig-Holstein, Campus Kiel, 24105, Kiel, Germany. **SYNOPSIS** Lymphocytic colitis (LC) is a non-destructive inflammatory bowel disease that displays a massive immune cell recruitment into the mucosa, probably less capable to respond than in its counterpart collagenous colitis (CC). Pathomechanisms leading to watery diarrhoea involve both paracellular and transcellular permeability of the apparently normal epithelial barrier and differ between LC and CC.

**Keywords:** Cell adhesion, inflammatory bowel disease, microscopic colitis, micro-RNA, RNA sequencing

## Abstract

**Background and Aims:** The pathobiology of the non-destructive inflammatory bowel disease (IBD) lymphocytic colitis (LC) is poorly understood. Our aim was to define a LC-specific transcriptome to gain insight into LC pathology, identify genetic signatures uniquely linked to LC, and uncover potentially druggable disease pathways.

**Methods:** We performed whole mucosa bulk RNA-sequencing of LC and CC samples from patients with active disease, and healthy controls (n=4-10 per cohort). Differential gene expression was analyzed by gene-set enrichment and deconvolution analyses to identify pathologically relevant pathways and cells, respectively, altered in LC. Key findings were validated using reverse transcription quantitative PCR and/or immunohistochemistry. Finally, we compared our sequencing data to a previous cohort of ulcerative colitis and Crohn’s disease patients (n=4 per group) to distinguish non-destructive from classic IBD.

**Results:** The LC-specific transcriptome was defined by a limited mucosal immune response against microbiota compared to CC and classic IBD samples. In contrast, we noted a distinct induction of regulatory non-coding RNA species in LC samples. Moreover, compared to CC, we observed decreased water channel and cell adhesion molecule gene expression, which was associated with reduced intestinal epithelial cell proliferation.

**Conclusions:** We conclude that LC is a pathomechanistically distinct disease that is characterized by a dampened immune response despite massive mucosal immune cell infiltration. Our results point to regulatory micro-RNAs as a potential disease-specific feature that may be amenable to therapeutic intervention.

## INTRODUCTION

Lymphocytic colitis (LC) is a debilitating inflammatory bowel disease (IBD) of the large intestine characterized by persistent non-bloody watery diarrhoea despite a macroscopically normal mucosa. The histopathologic hallmark of LC is an increased intraepithelial lymphocytosis (> 20 intraepithelial lymphocytes per 100 enterocytes), together with an inflammatory infiltrate – mainly lymphocytes and plasma cells with few eosinophils and neutrophils – in the lamina propria.^1–3^ The superficial epithelium can display focal damage including flattening, mucin depletion and vacuolization with occasional Paneth cell hyperplasia.^2^ LC incidence is estimated to be between 4.0-6.1 cases per 100,000 persons in Europe and the USA, with potentially increasing rates.^2,4^ LC is more frequent in older women (> 60 years old) but can occasionally affect younger individuals, including children.^2,4,5^ Based on guidelines, the standard first-line treatment to induce remission is oral corticosteroid budesonide, which is effective in 88% of patients.^1–3^ Still, disease often relapses after budesonide discontinuation.^6^

Dense genotyping has failed to identify genes associated with LC despite the mucosal immune cell infiltration and co-occurrence with the autoimmune disorder celiac disease.^7–9^ On the other hand, its counterpart collagenous colitis (CC) – collectively referred to as microscopic colitis –, has been associated with several human leukocyte antigen (*HLA*) genetic variants, which was confirmed by enrichment of adaptive immune transcriptional signatures.^7,8,10^ Besides clinical overlaps with CC, LC shares features with IBDs ulcerative colitis (UC) and Crohn’s disease (CD) such as Th1/Th17-driven immune responses, tight-junction claudin dysregulation, and water channel aquaporin downregulation.^11–19^ Interestingly, incidence of CD and UC increases in patients previously diagnosed of LC and CC.^6,20,21^ Indeed, both LC and CC have been suggested to be attenuated IBD forms where an overt mucosal inflammation remains restrained.^10,14,22^ Still, microscopic colitis can progress to classic IBD upon immune system hyperactivation as shown in LC patients overexpressing higher levels of interferon (IFN)- γ, and the Th2 transcription factor GATA3; or CC patients overexpressing tumor necrosis factor (TNF)-α and the Th1 transcription factor T-BET.^22^ Furthermore, LC displays mild and focal crypt architectural distortions (branched, dilated and disorganized crypts) and scattered non-necrotizing epithelioid granulomas and abscesses, similar to those observed in initial presentations of CD.^21,23,24^ Still, the extent of mucosal dysfunction and immune responses contributing to LC pathophysiology are unclear.

Water and sodium malabsorption have been suggested as pathomechanisms leading to diarrhoea in CC.^25–27^ However, epithelial surface injury is often more prominent in CC than in LC.^6^ Thus, mechanisms leading to diarrhoea in LC remain obscure and potential biomarkers or druggable targets are yet unknown. To address these questions, we investigated the whole transcriptome of colonic mucosa from LC patient samples and compared it to those of CC and classic IBD patients. Our patient cohort has enabled us to classify LC immune response as moderate, identify LC diarrhoea driving pathomechanisms, and propose targets for the development of new treatments for LC patients.

## RESULTS

### Lymphocytic colitis immune response is moderate

Lymphocytic colitis is characterized by chronic non-bloody diarrhoea and intact mucosa despite increased lymphocyte infiltration in both epithelia and lamina propria (figure 1A). Since LC pathophysiology is underexplored, we investigated its transcriptome, and compared it to that of healthy controls (Hc) and the closely related disorder CC. Principal component analysis grouped both LC and CC disease sample groups close to each other, with principal component 1 distinguishing them from healthy controls (figure 1B). LC mucosa displayed 5045 differentially expressed genes (DEGs) compared to healthy controls. These DEGs were mainly related to an inflammatory response and ion transport dysregulation (figure 1C-E, suppl. fig. 1). Leading gene-set enrichment terms included response to microbial-derived stressors such as lipopolysaccharide (LPS) involving both innate and adaptive immunity (antigen presentation through MHC complexes), resulting in chemoattraction, cytokine and humoral activity (figures 1C-E, suppl. Fig 1). Still, only 218 of the 5045 DEGs (4.32%) displayed a fold-change > |2|, indicating that these changes – despite statistically significant – might be modest compared to Hc (figure 1E). Our analyses also detected that DEGs (compared to Hc) could be targets of microRNAs (miR) 193b-3p, 615−3p, 192−5p, 215−5p, 92a−3p, and 16−5p (suppl. fig. 1).

**Figure 1.**
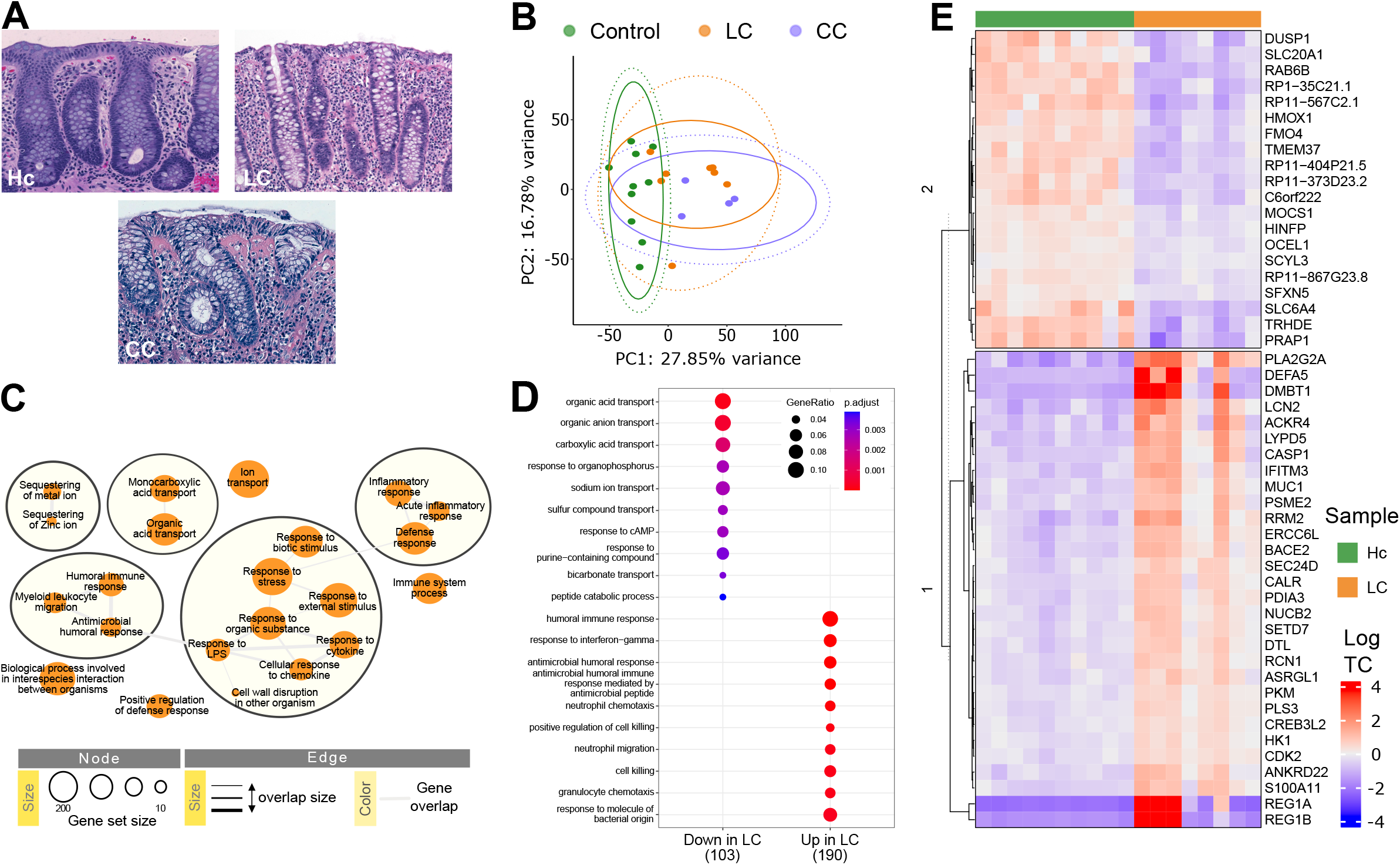
Lymphocytic colitis mucosal gene expression indicates activation of the central immune response. (A) Representative histology of hematoxylin and eosin–stained paraffin-embedded sections of the human colonic mucosa in a healthy control (Hc) subject, a lymphocytic colitis (LC) patient, and a collagenous colitis (CC) patient. (B) Principal component analysis of the RNA-seq expression profiles of LC (orange), CC (blue), and Hc (green). Ellipses indicate the multivariate t- (line) and normal (dotted line) distribution of gene expression profiles for each group with a confidence of 0.95. (C-D) Summarized enrichment map (C) and dot plot (D) of enriched biological process gene ontology in mucosal LC compared to Hc. The dot size in D indicated gene ratio as the ratio of significant differentially expressed genes to annotated genes in each gene set term. The colors display adjusted p-values. (E) Heatmap showing normalized log2-transformed fold changes (regularized log function in R) of the leading 50 differentially expressed genes (DEGs) between LC and Hc samples. n = 8–10 samples per group. Hc subjects are shown in green and LC samples in orange. Heatmap columns are split according to hierarchical clustering.

Compared to the clinically related disorder CC, LC differed in the expression of 384 genes (table 1, fig. 2A), of which 49 belonged to immunoglobulin (IG) genes (IGH, IGHV, IGK, IGKV, IGL or IGLV genes). Interestingly, all IG genes displayed a lower gene expression in LC compared to CC (median log_2_ fold change of -2.52, range -4.35 to -1.74) indicating a less prominent humoral immune response in LC (fig. 2B). Gene-set enrichment analyses also pointed to a less prominent cellular immune response in LC as pathways related to chemokine and cytokine activity displayed a diminished gene expression compared to CC (fig. 2C). Still, expression of CXCL16, CCL2, CCL3 and CCL11 in LC mucosa was diminished compared to CC (fig. 2D). To estimate immune cell type abundances, we imputed gene expression profiles and deconvoluted RNA-seq data (fig. 2E). Only naïve CD4+ T-cells were found significantly underrepresented in LC compared to Hc samples and were comparable to the estimation from CC samples (fig. 2E-F).

**Table 1.** Clinical and demographic characteristics of the patient cohort and controls.

| Variable | Hc | LC | CC |
| --- | --- | --- | --- |
| Total number of subjects | 10 | 8 | 4 |
| Gender, % females | 60.00 | 62.50 | 75.00 |
| Average age (range) | 57 (55-63) | 53 (20-80) | 70 (49-86) |
| Average stools/day (range) | - | 7.13 (2-15) | 7.50 (6-10) |
| Average watery stools/day (range) | - | 7 (2-15) | 7.50 (6-10) |
| Average collagenous band, μm (range) | - | - | 34.00<br>(12-52) |

CC, collagenous colitis; Hc, healthy controls; LC, lymphocytic colitis.

**Figure 2.**
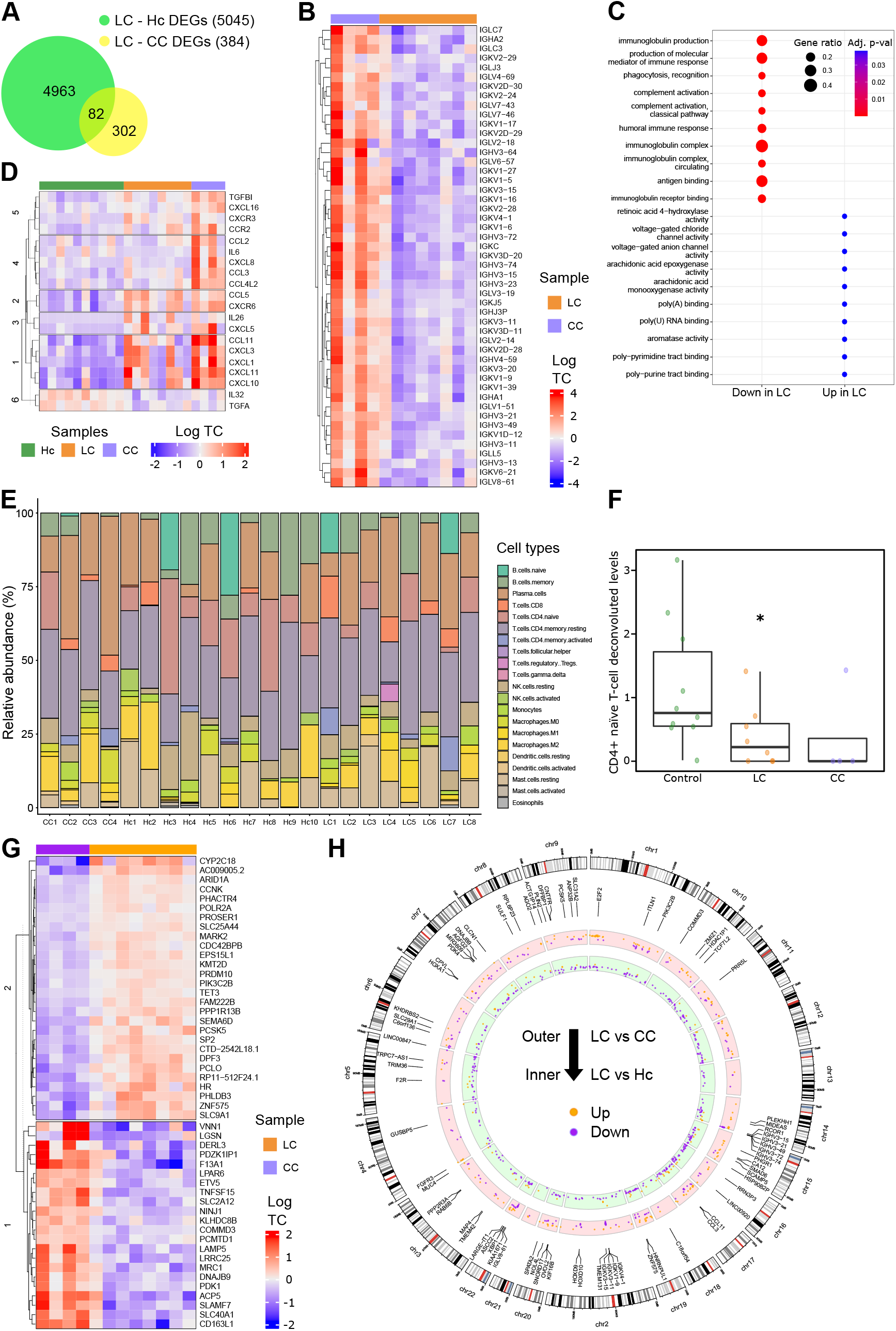
The immune response in lymphocytic colitis is limited compared to collagenous colitis. (A) Diagram displaying the numbers of differentially expressed genes (DEG) between lymphocytic colitis (LC), collagenous colitis (CC) and healthy control (Hc) sample groups. 5045 DEGs were found between LC and Hc samples (green), while 384 DEGs were identified between CC and Hc samples (yellow). A total of 82 DEGs overlapped between the two comparisons. (B) Heatmap displaying normalized log2-transformed expression values (regularized log function in R, log TC) of the immunoglobulin (IG)-related DEGs between LC and CC samples. (C) Dot plot of enriched biological process gene ontology in LC compared to CC. The dot size indicates gene ratio as the ratio of significant DEGs to annotated genes in each gene set term. The colors display adjusted p-values. (D) Heatmap displaying log TC of cytokine and chemokine DEGs between Hc, LC, and CC samples. (E) RNA-seq data deconvolution computed for different immune cells displaying the estimated relative percentage in each sample. (F) Plot of estimated immune cells according to RNA-seq data deconvolution that were statistically significant in LC compared to Hc samples (only CD4^+^ naïve T-cells). Median with interquartile range is shown. Statistically significant differences are shown as * p < 0.05. (G) Heatmap displaying log TC of the 50 leading DEGs between LC and CC samples, excluding IG-related DEGs. (H) Circos plot showing chromosomal distribution of leading DEGs upregulated (orange) or downregulated (purple) in LC compared to CC (outer plot) and Hc (inner plot) sample groups. Chromosome location and genetic distance (in Mb) are indicated in the outer most part of the plot. All results are based on n = 4–10 samples per group. LC subjects are shown in orange, CC samples in blue, and Hc in green. Heatmap columns are split according to hierarchical clustering.

Taken together, our results indicate that LC results from an immune response to luminal microbiota-derived antigens, which is less developed than the inflammation in CC. Therefore, despite the increased lymphocyte recruitment into LC mucosa, immune cells might remain relatively inactive to exert macroscopic mucosal damage.

### Lymphocytic colitis displays a general downregulation of water channel aquaporins and tight-junction claudins, which differ from the pattern in collagenous colitis

LC featured an enrichment of pathways related to the activity of voltage-gated anion channels, incl. chloride channels, arachidonic acid mono- and epoxygenases, and retinoic acid 4-hydroxylase (fig. 2C), which could be mechanisms contributing to diarrhoea in LC. The leading DEGs on these pathways included the chloride voltage-gated channel 1 (*CLCN1*) and the retinoid acid monooxygenase cytochrome P450 family 2 subfamily C member 18 or mephenytoin 4-hydrolase (*CYP2C18*) (fig. 2G-H). Interestingly, the gene for solute carrier family 9 member A1/Na^+^/H^+^ exchanger 1 (*SLC9A1/NHE1*), which is decreased in IBD,^28^ is upregulated in our LC cohort (fig. 2G).

Since water channel aquaporin (AQP) downregulation has been associated with IBD^18,29,30^ and was identified as a possible mechanism driving water malabsorption in the non-destructive IBD CC (AQP8 downregulation)^25^, we next aimed to evaluate AQP gene expression in our LC cohort (fig. 3). RNA-seq results showed a significant downregulation of *AQP7* and *AQP8* that was validated using RT-qPCR (fig. 3A-B). In addition, RT-qPCR identified a notable downregulation of many other AQP genes, including *AQP0-4, AQP6, AQP10, and AQP12* (fig. 3B). To further explore alternative mechanisms of paracellular permeability, we asked whether claudins (CLDN) and tight-junction protein 1 (*TJP1*) genes were also dysregulated. Among the most relevant claudins for intestinal biology, RNA-seq counts pointed to an upregulation of *CLDN1*, and the “leaky” *CLDN2*, and a downregulation of *TJP1* (fig. 3A). RT-qPCR validated the downregulation of *TJP1* only, and identified a significant downregulation of several claudin genes, including *CLDN3*, the “tight” *CLDN4, CLDN5, CLDN7, CLDN8*, and *CLDN15* (fig. 3D).

**Figure 3.**
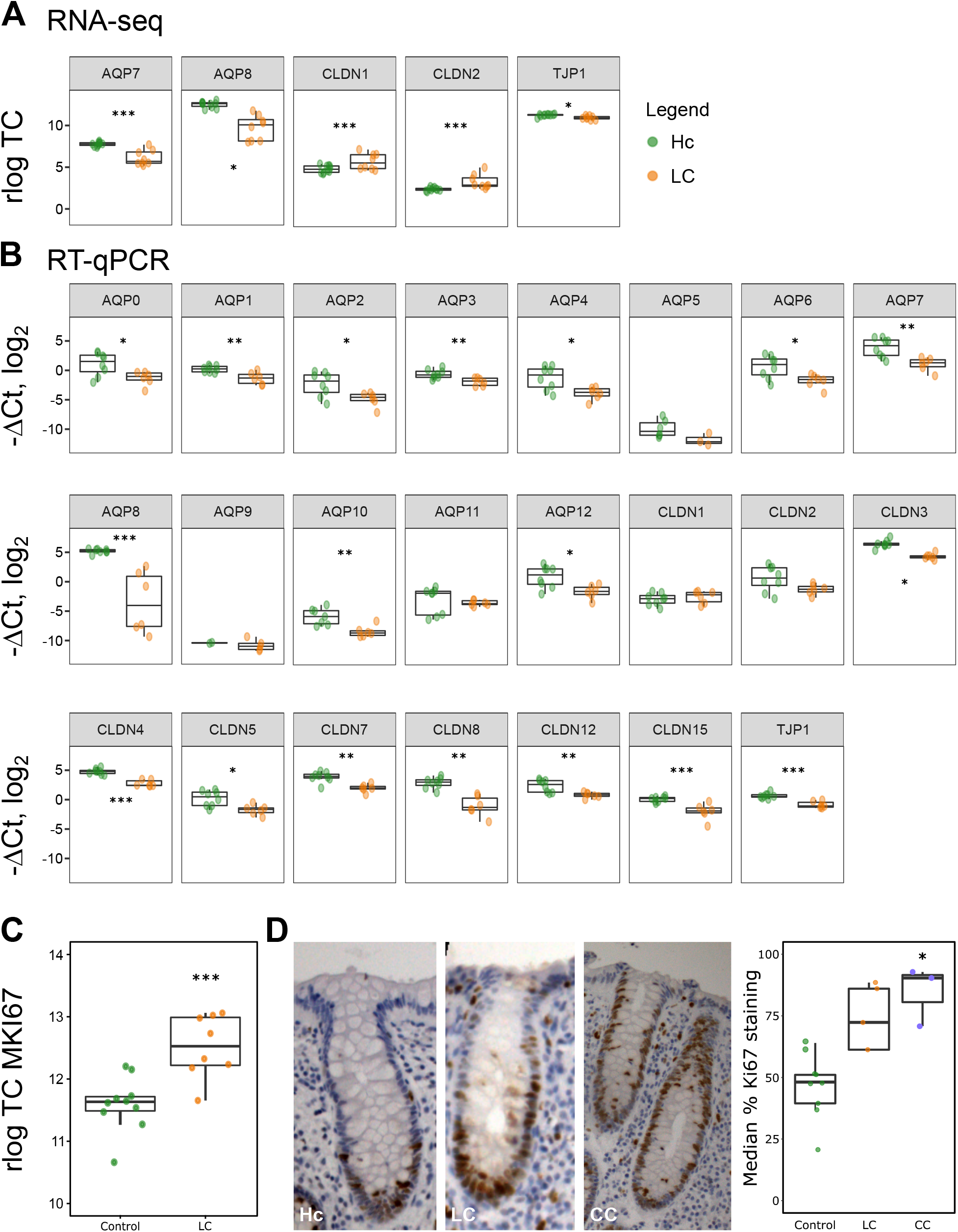
Epithelial cell homeostasis is disrupted in lymphocytic colitis. (A-B) Normalized log2-transformed expression values (regularized log function in R) of RNA-seq transcript counts (rlog TC, panel A) or log_2_ fold changes (–ΔCt, log_2_ values) in gene expression as analyzed by RT-qPCR (panel B) of water channel aquaporins (AQP), claudins (CLDN) and tight-junction protein 1 (TJP1) genes between lymphocytic colitis (LC, orange) and healthy control (Hc, green) samples. Median with interquartile range is shown for n = 6–10 samples per group. HPRT1 was used as a housekeeping control gene for RT-qPCR analyses. All primers detect all coding transcript variants of the indicated gene. (C) Log TC of MKI67 RNA-seq transcript counts. (D) Representative IHC images of longitudinally sectioned epithelial glands stained for Ki67 proliferation marker (brown) in paraffin-embedded sections of Hc, LC and collagenous colitis (CC) colonic mucosa (left). Analysis of Ki67 relative staining to total crypt length is shown on the right (median with interquartile range, n=3-9 well-oriented crypts per group, median of 8 crypts/patient). LC samples are shown in orange, CC in blue, and Hc in green. Statistically significant differences are shown as * p < 0.05, ** p < 0.01, and *** p < 0.001.

We previously described an intestinal epithelial hyperproliferation in colonic crypts from the mucosa of active CC patients.^10^ Since the expression of the proliferation marker MKI67 was upregulated in LC compared to healthy controls (fig. 3E, left panel), we next aimed to quantify the extent of the staining of Ki67 protein in LC. Our analysis showed that epithelial cells in LC samples proliferate at an increased rate compared to healthy controls, but in contrast to CC samples, this comparison was not statistically significant (fig. 3E). Collectively, these results point to an epithelial cell dysfunction where cells proliferate at relatively high rates and display altered paracellular and transcellular water transport.

### Posttranscriptional regulatory mechanisms may dampen the immune response in lymphocytic colitis

Classic IBD forms are distinguished by an overt immune response against microbiota that damages the intestinal tissue; however, LC mucosa is macroscopically intact.^21^ Thus, we next compared LC transcriptome data to classic IBD by using a patient cohort we previously published in Häsler R. *et al*.^31^ LC differed from active colonic CD in 4278 DEGs, and from active UC in 4740 DEGs, of which 60-80% were also differentially expressed between CD or UC and healthy controls. As expected, both CD and UC displayed a stronger immune response compared to LC, and, consequently, also exhibited reduced expression of genes associated with control over cell migration through blood vessels, angiogenesis, and endothelial cell proliferation processes (fig. 4A). Interestingly, regulatory transcriptional and translational mechanisms were enriched in LC samples, which indicated a role for miRNAs (fig. 4A-B). Enrichment analyses of DEGs for primary transcripts of miRNAs highlighted those involved in extracellular matrix and transmembrane receptors that mediate cell-to-cell communication (fig. 4C). Gene-set enrichment analyses of DEGs that were unique for each of the comparisons additionally indicated increased cytokine activity in UC compared to LC and decreased sensory perception of chemical stimuli in CD compared to LC (fig. 4D).

**Figure 4.**
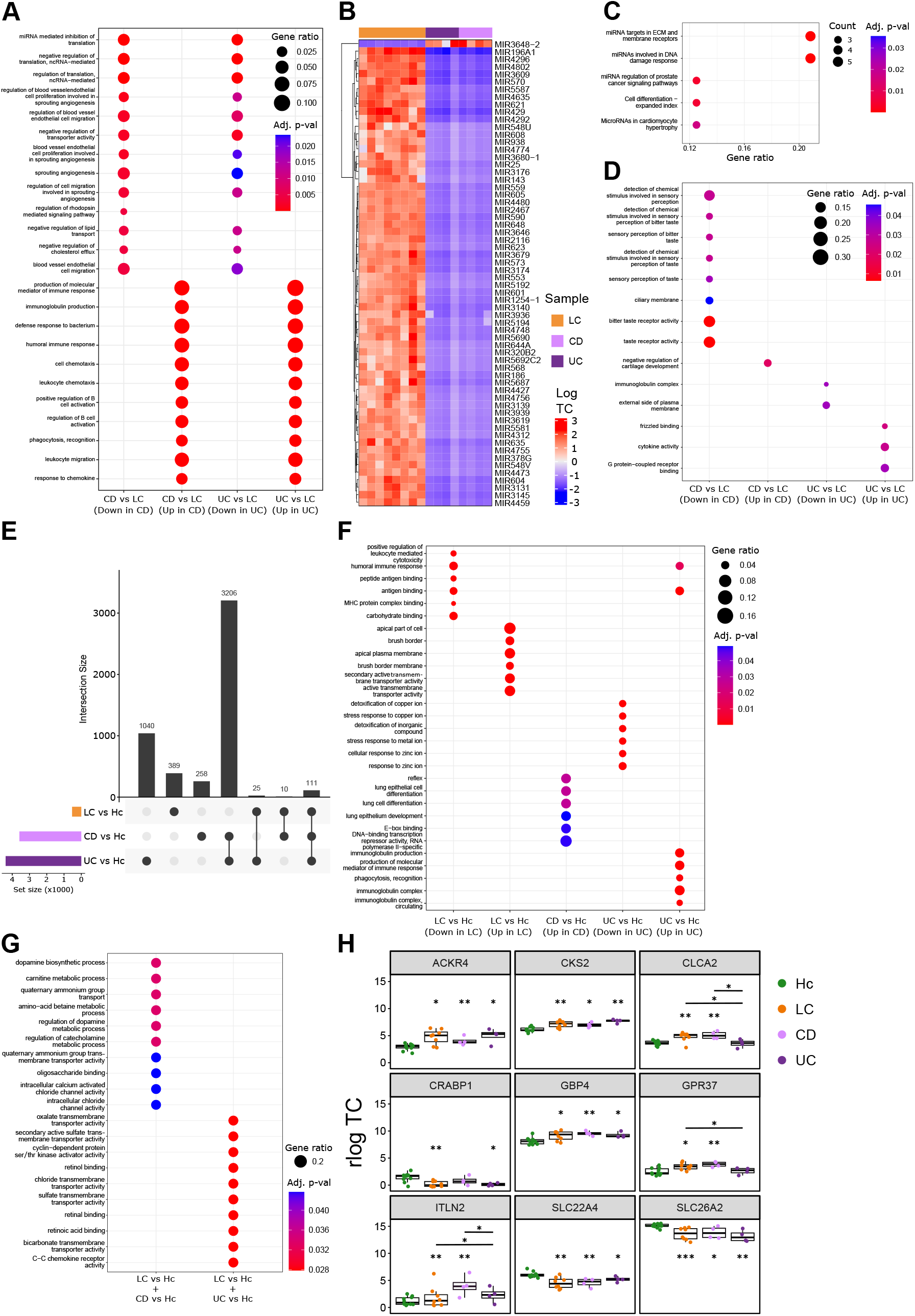
Lymphocytic colitis immune response is decreased compared to classic IBD forms and might be restrained by posttranscriptional regulatory mechanisms. (A) Dot plot of enriched biological process gene ontology (GO:BP) in LC compared to Crohn’s disease (CD) and ulcerative colitis (UC). (B) Heatmap displaying normalized log2-transformed expression values (regularized log function in R, log TC) of leading differentially expressed genes (DEG, log_2_ fold-change > |5|) of primary precursors of micro-RNAs (MIR) between LC, CD and UC samples. Heatmap columns are split according to hierarchical clustering. (C) Dot plot of leading enriched primary precursor MIR DEGs according to WikiPathway database. (D) Dot plot of enriched GO:BP in LC compared to CD and UC groups, including only unique DEGs in the comparison of LC with CD, and of LC with UC. (E) Intersection plot of DEGs from comparisons of disease (LC, CD, or UC) samples against healthy control (Hc) samples. (F) Dot plot of enriched gene ontology between disease (LC, CD, or UC) compared to Hc, including only unique DEGs for each of the comparisons. (G) Dot plot of enriched gene ontology for shared DEGs between LC or CD comparisons to Hc (n=10), or between LC or UC comparisons to Hc (n=25). (H) Normalized log2-transformed expression values (regularized log function in R) of RNA-seq transcript counts in gene expression of leading genes from the dot plot in G between LC, CD, UC and Hc samples. All results are based on n = 4-10 samples per group. LC subjects are shown in orange, CD samples in light purple, and UC samples in dark purple. Dot sizes in enrichment dot plots indicate gene ratio as the ratio of significant differentially expressed genes to annotated genes in each gene set term, while the colors display adjusted p-values. Statistically significant differences are shown as * p < 0.05, ** p < 0.01, and *** p < 0.001 against Hc samples unless indicated otherwise.

To further explore the similarities and differences with classic IBDs, we used the comparisons of each disease with healthy controls. Among the unique DEGs for each of them, brush border remodeling and apical transport pathways were upregulated in LC compared to Hc, and epithelial differentiation was upregulated in CD (fig. 4F). On the other side, UC displayed a unique upregulation of pathways related to the immunoglobulin production and phagocytosis, and downregulation of detoxification events involving copper and zinc (fig. 4F). When intersected comparisons were analyzed, LC shared with CD the activity related to amino acid, dopamine, and catecholamine metabolic processes, and calcium activated chloride channels; and LC shared with UC retinoic acid binding, and transmembrane transport activities (fig. 4E, G). However, the DEGs leading to these pathways were limited (fig. 4H). To note, gene expression of oligosaccharide binding intelectin *ITLN2* could distinguish CD from LC and UC, and calcium-activated chloride channel *CLCA2* and the G protein-coupled receptor GPR37 could distinguish UC from LC and CD (fig. 4H). In summary, LC immune response, in contrast to classic IBD, could be restrained by miRNA-driven regulatory events that affect cell-to-cell contact and extracellular matrix remodeling, which could be exploited for the development of new treatments for IBD patients.

## DISCUSSION

Lymphocytic colitis aetiology and pathophysiology are poorly understood. Here, we describe a transcriptional alteration of genes related to antigen presentation, including lipopolysaccharide response, that point towards a moderate immune response in LC when compared to its counterpart CC.

Even though microscopic colitis, including LC, has been associated with autoimmune disorders,^32^ dense genotyping studies discarded any link of human leukocyte antigen (HLA) genes with LC.^7,8^ A report shows an increase of HLA-A1 and a decrease of HLA-A3 in LC;^33^ however, we found no indications of such due to the limited expression of HLA molecules (only *HLA-G* expression increased when compared to Hc with a log2 FC of 2.95, adjusted p-value = 0.02). Instead, we found an activation of humoral immune responses as IG-related gene expression was induced. This result matches the observation that anti-nuclear antibodies (ANA), anti-Saccharomyces cerevisiae antibodies (ASCA) IgG, and thyroid peroxidase (TPO) antibodies are more prevalent in LC than in controls.^34^ Interestingly, Epstein-Barr virus is nearly always detectable in microscopic colitis biopsies so it could explain the increased viral immune response we found in LC – and previously reported in CC^10^ –, which link both disease entities with autoimmunity due to its ability to mimic host molecules.^35,36^

We have also reported an active cellular immune response in LC, but to a lesser extent than CC, CD and UC. Günaltay *et al*. reported an increased expression of granulocyte, Th1 and CD8^+^ T cell-associated chemokines in LC mucosa^16^ that we partially corroborate. However, we found that expression levels of immunoglobulin-related genes and key chemoattractants such as CXCL8/IL8 was lower in LC compared to CC. Going forward, our deconvolution analysis pointed to a decrease in CD4^+^ naïve T-cells in LC which was comparable to that on CC samples. In contrast, Carrasco *et al*. reported more robust results using flow cytometry, demonstrating an increase in CD3^+^ T-cells, CD3^+^CD8^+^ T-cells, CD3^+^CD4^+^TCRγδ^+^ T-cells, double negative CD3^+^CD4^-^CD8^-^ T-cells, and a decrease in CD3^+^CD4^+^IFN^+^ Th1 and CD3^+^CD4^+^IL-17A^+^ Th17 cells in LC.^14^ Despite of lower Th1/Th17 cell numbers, *IFNG, TNFA, IL17A, IL21, IL23* gene expression levels were increased, which were also reported by Kumawat *et al*.^41,42^ We can only corroborate the increase of *IFNG* gene expression (log_2_ FC = 5.18 vs Hc) probably due to individual variability and the use of different cohorts, but more interesting, none of these reports find a clear association between cytokine gene expression and protein levels. Therefore, LC comprises a disorder where immune cells are intensively recruited to the tissue. Whether these cells have limited activity and remain as already recruited ready-to-react sentinels remains unknown.

The extensive water channel AQP downregulation we found in LC contrasts the unique downregulation of AQP8 in CC.^25^ In addition, tight-junction claudin gene expression is downregulated, indicating a combined paracellular and transcellular permeability in LC. Furthermore, the increased expression of calcium channel *CLCN1* and sodium-hydrogen exchanger *SLC9A1/NHE1*, and limited expression of epithelial sodium channel (ENaC) γ subunit^37^ could contribute to chloride and sodium disbalance in LC, exacerbating water malabsorption in the colon, hence, leading to diarrhoea. On the other side, downregulation of sodium-hydrogen exchangers SLC9A2/3 (NHE2/3), downregulated in adenoma (SLC26A3/DRA), and putative anion transporter 1 (SLC36A1/PAT1) seem to be responsible for misbalanced ion exchange in CC.^25,38,39^ Altogether, these results point to an increased water and ion permeability in LC, and differ from the diarrhoeal mechanisms described for CC. Still, whether ion channels and tight-junction leakage initiate the mucosal exposure to luminal insults in LC or are a consequence of pre-activated immune response in the highly proliferative epithelia and underlying immune cells remains unknown.

When we compared LC to classic IBD forms, we found that miRNAs could have a role controlling an overt immune response and contributing to tissue remodeling in LC. However, the increased expression of primary transcripts of miRNAs we found in LC when compared to CD and UC could be an artifact of comparing datasets from different sequencing batches. Since the only miRNA study to date with sufficient microscopic colitis samples found that the CD phenotype-linked stress responsive miR-31 was upregulated in IBD and CC, but not in LC^40,41^, further studies and validation are required to describe the role of miRNAs in LC.

In summary, our results confirm that LC is an immune-mediated IBD in which a limited immune response against luminal antigens induce extensive immune cell recruitment. In comparison to CC, CD and UC, LC immune response is mild and could be restricted by miRNA modulation. Diarrhoeal mechanisms in LC are distinct as water channel and tight-junction claudin downregulation is generalized, compared to the exclusive AQP8 downregulation in its counterpart CC. Also, ion channels involved in LC pathogenesis (CLCN1, NHE1, ENaCγ) differ from those in CC (NHE2/3, DRA, PAT1). This, together with an increased proliferative rate of intestinal epithelia, demonstrates an epithelial cell dysfunction in LC.

## MATERIALS AND METHODS

### Study population

Biopsy samples from the descending colon were collected during scheduled colonoscopy in adult patients with lymphocytic colitis (LC) and collagenous colitis (CC) patients at the Division of Gastroenterology at Linköping University Hospital, Sweden. LC and CC were diagnosed according to the current guidelines.^2^ LC diagnose is primarily based on clinical history and histopathological features, including intraepithelial lymphocytosis (≥20 lymphocytes per 100 surface epithelial cells), an increased inflammatory infiltrate in lamina propria and a not significantly thickened collagenous band (<10 μm). CC diagnose relies on clinical history and the histopathological finding of a subepithelial collagen band of > 10 µm thickness. Active CC and LC were defined as more than three bowel movements/day or at least one watery bowel movement/day during a one-week registration period.^2,42^ Healthy volunteers were recruited from the local colon cancer screening program at Linköping University Hospital (Sweden). These individuals showed normal macro- and microscopic findings upon histopathological assessment, had normal bowel movements, did not take any medication at the time of colonoscopy, and did not suffer from any gastrointestinal or autoimmune diseases. Detailed patient characteristics can be found in table 1. Adjacent biopsy samples from the same mucosal area were stored in AllProtect (Qiagen, Hilden, Germany) for subsequent RNA extraction, or in PBS for fixation in paraformaldehyde, embedding in paraffin and analyses using microscopy. Informed written consent was obtained from all subjects, and their data were handled according to current regulations (EU2016/679, corrigendum 23 May 2018). Ethical approval was issued by Linköping’s regional ethical committee to conduct studies in microscopic colitis, including CC and LC (Dnr 2015/31-31).

### Genome-wide mRNA sequencing (RNA-seq)

Biopsies preserved in AllProtect were homogenized in RLT buffer from RNeasy Mini Kit supplemented with 1% 2-mercaptoethanol in a TissueLyser II instrument (all from Qiagen, Hilden, Germany). Total RNA from homogenized biopsy samples was isolated using RNeasy Mini Kit (Qiagen) following the manufacturer’s instructions. RNA sequencing libraries were constructed with Illumina TruSeq Stranded total RNA RiboZero GOLD (Illumina, San Diego, CA), and paired-end sequenced (2 × 150 bp reads) for 300 cycles to a depth of 1,000 million base reads on a NovaSeq6000 instrument (Illumina) according to the manufacturer’s recommendations. FASTQ files were generated using bcl2fastq software v2.20. Data was analyzed using the R Bioconductor software v3.5.1 (R Core Team 2018), including SARTools v1.6.6 and DESeq2 v4.1.3 packages.^43–45^ Reads were aligned to the Ensembl GRCh38 genome version, release 92.

### RNA-seq data analysis

Differential gene expression (DGE) from RNA-seq data was determined with linear models using DESeq2 v4.1.3 and significance decided by Benjamini-Hochberg false discovery rate (FDR) adjusted p-values <0.05. Raw count data was normalized using DESeq2 with the regularized logarithmic function (rlog transformed expression values) in R for subsequent analyses and representation. Principal component analysis (PCA) was computed after making the data homocedastic using R. Differentially expressed genes (DEG) were filtered according to >|1.5| log_2_ fold-change for subsequent gene ontology (GO) biological process and Kyoto encyclopedia of genes and genomes (KEEG) enrichment analyses using clusterProfiler v4.2.2, and representation using dot plots. Intersection plots were created using VennDetail v1.10.0.

In addition, enrichment maps were generated after gene-set enrichment analyses (GSEA) using EnrichR taking into account GO, KEGG and WikiPathway databases after DEGs were filtered for genes with >10 counts and ranked according to their log_10_ adjusted-p-value and sign of log fold-change. EnrichR GSEA results for comparisons of two sample groups was performed in GSEA^46,47^ v4.0.3 (Broad Institute, Cambridge, MA) using updated guidelines.^48^ Human gene set annotations from GO, Reactome and other databases were downloaded from Bader’s Lab website excluding those inferred from electronic annotations (https://download.baderlab.org/EM_Genesets/, release November 2021). Terms annotating more than 200 or less than 10 genes were discarded to improve biological interpretation. GSEA results were visualized using EnrichmentMap^49^ v3.2.0 and nodes were generated using a FDR p-value of less than 0.05, a Jaccard Overlap Combined index of 0.375 and k constant of 0.5, and annotated using AutoAnnotate v1.3 applications for Cytoscape v3.7.2, and improved in Inkscape v0.92.4. Additionally, GSEA was complemented with the identification of potential microRNA-gene target interactions using miRTarBase database coupled to g:Profiler public web server.^50,51^

Chromosomal visualization of DEGs was performed using online shiniCircos server (http://shinycircos.ncpgr.cn).^52^ Deconvolution was computed using CIBERSORTx online tool (https://cibersortx.stanford.edu/)^53^ to impute gene expression profiles and estimate cell type abundances using RNA-seq data.

### Immunohistochemistry (IHC)

Paraffin-embedded sections (4 µm) were cut in a microtome and deparaffinated with Xylol substitute (Carl Roth GmbH, Karlsruhe, Germany). Antigen retrieval was performed in a pressure cooker with citric acid pH 6.0 for 20 min. Samples were incubated with 3% H_2_O_2_ for 10 min to block endogenous peroxidase activity and subsequently with 1% bovine serum albumin (BSA) in PBS with 0.2% TritonX100 for 30 min. Anti-Ki67 (dilution 1:500, 556003, BD Biosciences, Franklin Lakes, NJ, EE.UU.), and secondary goat anti-mouse IgG biotin conjugated antibody (ab6788, Abcam) were used to stain the samples in PBS with 1% BSA (Sigma-Aldrich). The VECTASTAIN Elite ABC reagent was used to develop the staining (Vector Laboratories, Burlingame, CA). Counterstaining with hematoxylin (Hemalum, Carl Roth) was performed before dehydration with ethanol series and mounting with with Roti Histokitt II mounting medium (Carl Roth, Karlsruhe, Germany) were carried out. Images were acquired on a Zeiss AxioImager.Z1 microscope (Zeiss, Jena, Germany).

### Reverse transcription quantitative polymerase chain reaction (RT-qPCR)

Total RNA from biopsies preserved in AllProtect were homogenized as mentioned above (see genome-wide mRNA-sequencing section). RNA was quantified using a NanoDrop ND-2000 and reverse-transcribed with a High Capacity cDNA Reverse Transcription Kit (all from Thermo Fisher Scientific). Relative gene expression was quantified by RT-PCR with iTaq Universal SYBR Green Supermix (BioRad, Hercules, CA) following the manufacturer’s instructions and using the primer pairs in supplementary table 1 besides the ones already reported.^25^ Primers were designed to amplify all transcript coding variants of the selected gene in the Reference Sequence (RefSeq) collection of the National Center for Biotechnology Information (NCBI, Bethesda, MD) taking the longest transcript sequence as a reference, with primers annealing in different exons for all transcript variants – when possible – using Primer3Plus v2.4.2 software.^54^ Quantitative analysis was carried out in a CFX96 Touch Real-Time PCR detection system (BioRad) using the relative quantification -ΔCt method. Hypoxanthine phosphoribosyltransferase (*HPRT*) *1* was used as a reference gene, and each sample was analyzed in duplicate.

### Statistical analyses

Ki67 median percentage of staining in crypts from human colonic samples were analyzed with the non-parametric Kruskal-Wallis test when different groups were compared among each other. Quantitative PCR data (-ΔCt values) were analyzed with the non-parametric Kruskal-Wallis test and Mann-Whitney test, and adjusted according to Benjamin-Hochberg FDR. Statistical analyses were performed and plotted in R.

All authors had access to the study data and reviewed and approved the final manuscript.

## Data Availability

All data produced in the present study are available upon reasonable request to the authors.

## ABBREVIATIONS

CC: collagenous colitis
CD: Crohn’s disese
DEGs: differentially expressed genes
DGE: differential gene expression
IBD: inflammatory bowel disease
LC: lymphocytic colitis
log TC: normalized log_2_ transformation of RNA-sequencing transcript counts
Log2FC: log_2_ fold-change
RNA-seq: RNA-sequencing
UC: ulcerative colitis

## Acknowledgements

We thank Lena Svensson (Linköping University) for support collecting human samples and Maren Reffelmann and Tanja Klostermeier (University Hospital Schleswig-Holstein) for support with IHC. We also thank the National Genomics Infrastructure in Genomics Production (NGI) in Stockholm, Sweden, where the gene expression analysis of RNA sequencing data was performed. NGI is funded by Science for Life Laboratory, the Knut and Alice Wallenberg Foundation and the Swedish Research Council, and SNIC/Uppsala Multidisciplinary Center for Advanced Computational Science for assistance with massively parallel sequencing and access to the UPPMAX computational infrastructure. We are grateful to all volunteer patients who agreed to participate in the study.

## AUTHORS’ CONTRIBUTIONS

C.E.H. designed, performed, and analyzed most experiments. A.B. contributed to RNA-seq analysis and J.M. contributed to IHC analyses. C.E.H., S.K. and A.M. conceived the study; and C.E.H., S.K., A.M. and P.R. supervised the study. A.M. enrolled and sampled microscopic colitis patients and volunteers that participated in the study. C.E.H. wrote the manuscript. All authors provided with critical revision of the manuscript, reviewed, and approved the final version of the manuscript.

## SUPPLEMENTARY FIGURE LEGENDS

**Supplementary figure 1.** LC mucosa gene expression indicates activation of the central immune response. (A-B) Complete enrichment map of mucosal lymphocytic colitis (LC) gene expression compared to healthy control (Hc) samples according to Gene Ontology (GO) Biological Process (A) and GO Molecular Function (B) databases. (C) Top 20 leading miRNA terms resulting from miRNA gene target analysis of DEGs in LC samples compared to Hc.

## SUPPLEMENTARY TABLES

**Supplementary table 1.**
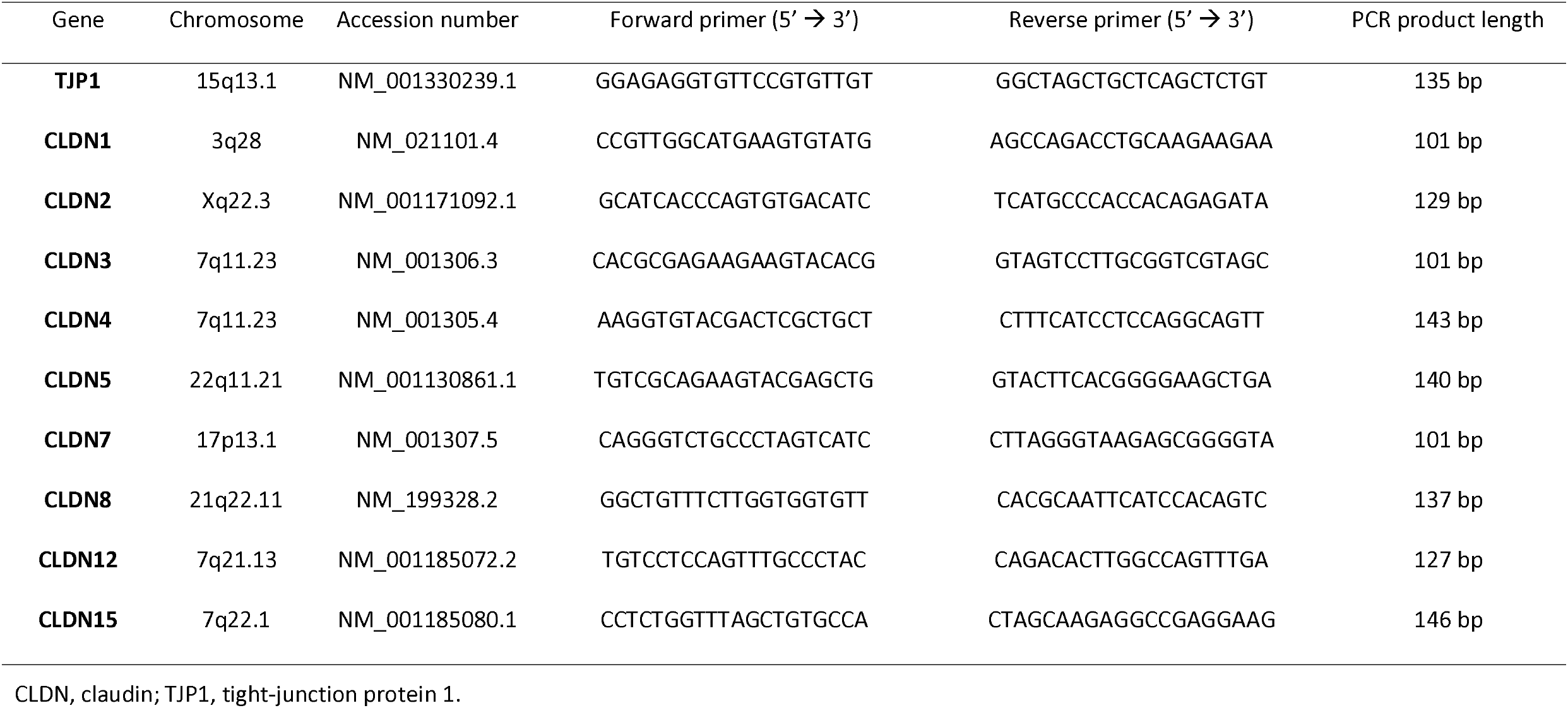
Sequence of the primer pairs used for quantitative PCR.

