## Supplementary figure 1 for "Transcriptomic profiling of lymphocytic colitis highlights distinct diarrhoeal pathomechanisms"

**A**

Complete GO Biological Process enrichment map

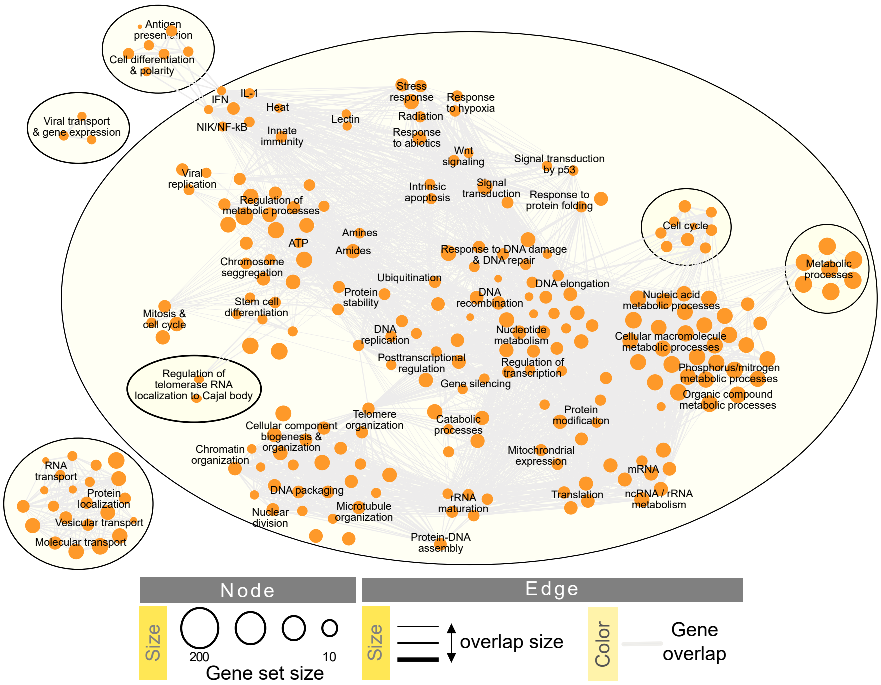


**B**

Complete GO Molecular Function enrichment map

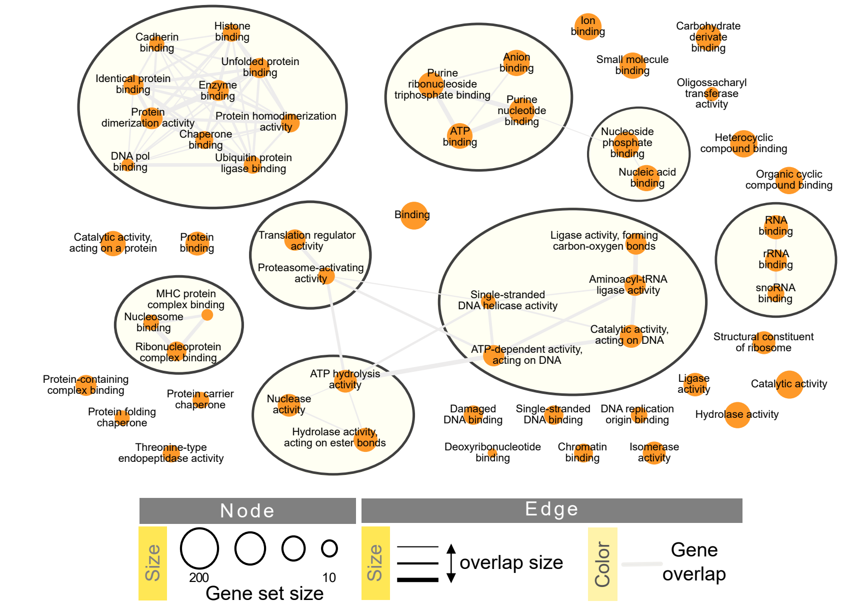


**C**

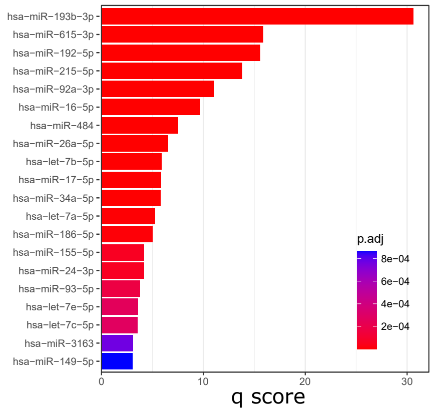
